# Respiratory virus genomic epidemiology during post-pandemic re-emergence of influenza in Australia

**DOI:** 10.1101/2025.02.23.25320087

**Authors:** Rebecca J Rockett, Jessica E Agius, Shona Chandra, Winkie Fong, Ammar Aziz, Carl JE Suster, Kerri Basile, Connie Lam, Sharon C-A Chen, Dominic E Dwyer, Jen Kok, Sheena G. Sullivan, Ian G. Barr, Vitali Sintchenko, Tanya Golubchik

## Abstract

Simultaneous genomic sequencing of multiple respiratory pathogens from clinical samples can provide real-time data on viral evolution, co-circulation and co-infection during seasonal epidemics. Influenza is a major global respiratory pathogen, with a mature genomic epidemiology framework, that has yet to be integrated with genomic surveillance of other co-circulating respiratory viruses. Leveraging existing well-integrated community influenza sampling during the first major Australian winter influenza season after the lifting of the COVID-19 pandemic restrictions in 2022, we examined the genomic epidemiology of influenza alongside co-infections of common human respiratory viruses. Using a commercial respiratory viral sequencing method, Respiratory Virus Oligo Panel (RVOP, Illumina), we recovered full-length human influenza A genomes from 75% (117/157) of samples with nucleic acid amplification test-confirmed influenza, as well as 19 genomes of co-infecting viruses from 17 samples, including respiratory syncytial virus (RSV-B), human bocavirus, SARS-CoV-2, human metapneumovirus, and coronaviruses 229E and OC43. The observed incidence of co-infecting viruses was temporally consistent with national epidemic trends (RSV and SARS-CoV-2). In all co-infections, the viral abundance was predominated by one of the infecting viruses, suggesting either consecutive infections, or within-host dynamics favouring the dominance of one of the infecting viruses. The dominant influenza subtype was A/H3N2, with a persistent minority of A/H1N1 infections, consistent with national surveillance. We contextualise our representative sample set within the global genomic diversity of A/H3N2 and the ongoing evolution of influenza A/H3N2 in the following Northern Hemisphere 2022/2023 winter. In addition to influenza genomic epidemiology, multi-pathogen methodology enables simultaneous detection and characterisation of co-infecting respiratory pathogens, providing insights into the role of viral dynamics during overlapping epidemics.

## Introduction

During the Southern Hemisphere 2022 winter, the seasonal community influenza virus epidemic coincided with epidemic waves of multiple respiratory viral infections, including SARS-CoV-2, respiratory syncytial virus (RSV) and human metapneumovirus (hMPV) (1). On a population scale, overlapping respiratory virus epidemics can rapidly escalate the demand on the healthcare systems, both via a direct increase in the total number of affected individuals, and indirectly, via the potential for more severe infections in the presence of multiple co-infecting pathogens (2, 3). The dynamics and complexities of respiratory co-infections and overlapping epidemics are only beginning to be appreciated, with important consequences for predicting disease severity, (4) understanding viral interference (5, 6) and uncovering novel pathogenesis due to pathogen-pathogen enhancement (7, 8).

The pivotal role of genomic epidemiology in real-time analysis of the COVID-19 pandemic has generated widespread expectations of similarly timely and high-resolution genomic monitoring of other circulating pathogens to support public health containment strategies, healthcare preparedness and vaccine design (9-11). Historically, influenza genomics capability was one of the earliest to be developed (12, 13), driven by the well-recognised pandemic potential of influenza A, which was responsible for four pandemics in the last century alone (H1N1 (1918), H2N2 (1957), H3N2 (1968) and H1N1 (2009) (13, 14)), as well as the current epidemic of highly pathogenic H5N1 avian influenza first identified in 1997(15). Even partial genomic information, such as sequences of the antigenically relevant haemagglutinin (HA) and neuraminidase (NA) genes, is vital for vaccine design and epidemic modelling. More recently, efforts have been made to develop genomic epidemiology for RSV (16-19), which are likely to be expanded with the introduction of new RSV vaccines and therapeutics. However, genomic surveillance of other respiratory viruses remains minimal.

A limitation of most viral genomics methodologies is that laboratory protocols are designed around a single pathogen. Co-infections are seldom tracked, and no single public health workflow can accommodate multi-pathogen (syndromic) genomics. As broader sequencing strategies begin to generate syndromic genomic data (9, 11), there is a strong incentive to design and evaluate workflows that can make use of these data to enhance epidemic management.

### Post-pandemic rise in influenza A in Australia

Epidemic dynamics of respiratory virus circulation are strongly influenced by pre-existing population immunity (20). The global implementation of stringent public health measures due to the COVID-19 pandemic limited the circulation of many respiratory viruses, including Influenza A virus (21-23). As restrictions eased and international travel resumed, Australia experienced an early peak of seasonal influenza in 2022 (24). Most local cases were diagnosed between May and June, with the highest case load occurring in the fortnight ending 26 June 2022 (25). Compared with the preceding 5-year average, the peak of infections occurred approximately three months earlier (May 2022) and resulted in a 3-fold higher rate of notifications to public health authorities. In total, the 2022 season reported 225,332 of laboratory-confirmed influenza cases, with 82.7% determined to be influenza A virus (25). The rapid increase in cases coincided with limited vaccine protection, due to the early rise in influenza infections prior to the usual delivery of seasonal influenza vaccinations and low uptake of influenza vaccines in the community. Only 1.4% of children aged less than five years and 12.4% of adults greater than 65 years were vaccinated by mid-April 2022 (26). This low vaccine uptake could be a result of vaccine fatigue and hesitancy, and barriers to seeking healthcare during the COVID-19 pandemic (27-29). A similarly early spike in influenza cases was also reported in Europe and North America during November and December 2022, the start of the Northern Hemisphere winter season (30).

The genomics of influenza A virus is crucial for providing data on the genetic diversity of circulating influenza viruses, when supported by phenotypic characterisation it enables an informed decision on candidate vaccine viruses for the following influenza season (31). In this study, the extent of genome-wide viral diversity among circulating influenza viruses in the 2022 season was assessed and identified epitope changes within the primary antigen of the dominant circulating influenza A subtype, A/H3N2, in addition to tracking corresponding rates of co-infections.

Here we explored a commercially available method for multi-pathogen targeted sequencing to simultaneously detect and reconstruct influenza A genomes alongside other common respiratory viruses, including viruses detected among patients co-infected with an influenza virus and another virus, in patient samples within a public health setting. Leveraging the existing well-integrated surveillance strategy for influenza in the community in Australia during the 2022 winter season, we used influenza-positive samples to analyse the genomic epidemiology of influenza in the context of other respiratory viruses, including RSV and SARS-CoV-2.

## Results

### Sample collection and viral genome sequencing

For this study, we retrieved remnant samples referred to the Institute of Clinical Pathology and Medical Research (ICPMR), NSW Health Pathology from collecting laboratories across the state of New South Wales (NSW) and the Australian Capital Territory (ACT). Together NSW and ACT make up 34% of the Australian population, which includes Sydney, a city of over 6 million people and the most common point of entry to Australia for international arrivals during the study period. Samples were retrieved in near-to-real time throughout the 2022 influenza season and sequenced in batches every 2-4 weeks. From 25 March to 14 July 2022, 180 unique samples positive for influenza on routine RT-PCR had sufficient residual extract to be sequenced as part of this study (Supplementary Figure S1). The majority (71%, 127/180) were from symptomatic individuals within the community, a quarter were hospital inpatients (26%, 46/180) and 3% were from high dependency or intensive care units. The cohort contained both children (>18 years, 44%, 80/180) and adults (55% 100/180) (Supplementary Figure 3).

It is notable that sequencing was done from remnant samples, which introduces an inevitable freeze-thaw cycle, and this may consequently reduce RNA quality. On re-testing, 23/180 or 12.7% of specimens were negative for influenza by RT-PCR. We included all 180 samples in our sequencing workflow but, as expected, influenza virus reads could not be recovered from the PCR-negative samples.

Using the RVOP enrichment protocol with Illumina sequencing, 130/157 PCR-confirmed samples (83%) yielded sufficient influenza A sequence reads to infer subtype, and of these, 117/130 (90%) produced a high-quality consensus influenza A virus genome (Supplementary Figure 1). Reconstructed genomes had a median depth of 682x and median genome coverage of 13,161-bp, with complete coverage of all eight influenza A genomic segments (Supplementary Table S1). Successful genome reconstruction was highly concordant with the quantity of influenza RNA template, consistent with previous reports that targeted capture protocols are quantitative with respect to viral load (18, 23, 32, 33). Specimens from which a complete genome was obtained had mean cycle threshold (Ct) value of 22.5 [interquartile range 21-24], whereas specimens from which we could not reconstruct whole genomes had mean Ct of 30.2 [interquartile range 26-32], including the 23 specimens where influenza virus could not be detected despite repeat RT-PCR testing, consistent with RNA degradation (Supplementary Figure 1).

### Spatiotemporal viral dynamics

Among the sequenced cohort, the temporal distribution of cases followed trends of influenza notifications at both state and national level, with the peak of cases in the fortnight ending 26 June 2022 (Figure 1). The influenza A/H3N2 subtype predominated across all timepoints after 10 April 2022, which also reflected state and national reports of A/H3N2 dominance. Concurrently, there was an ongoing wave of SARS-CoV-2 infection, dominated at the time by Omicron variant sublineage BA.2, and a rapidly developing RSV epidemic (Figure 1). Over the peak influenza period, the week ending 25 June 2022, 55,510 COVID-19 cases, 13,409 laboratory-confirmed influenza cases, and 2,700 RSV laboratory-confirmed cases were reported in NSW, with a combined incidence more than double the five-year average of emergency department presentations for ‘influenza-like illness’ (34).

**Figure 1.**
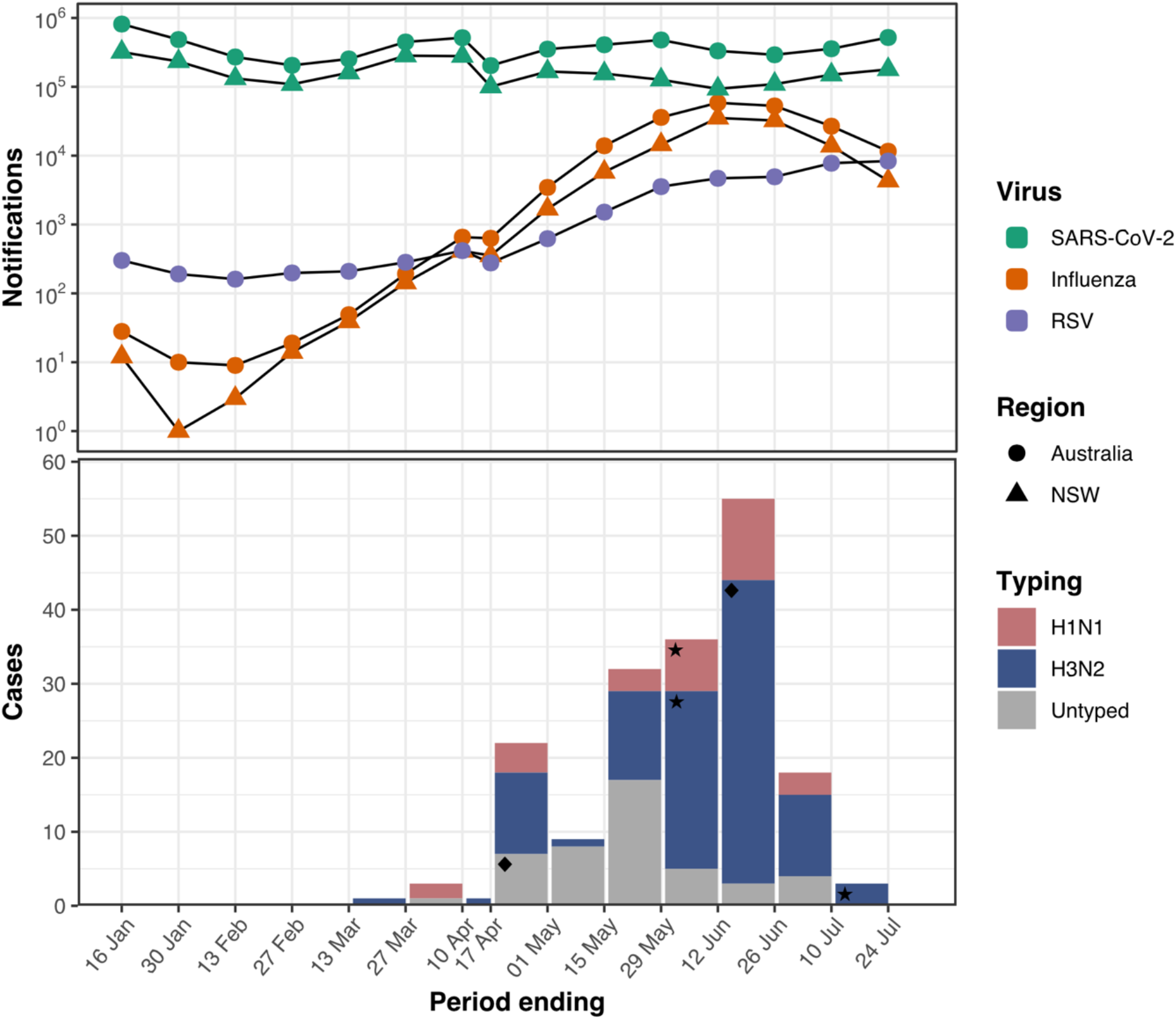
Co-circulation rates of human Influenza, RSV and COVID-19 infections in NSW and Australia during the study period **Figure 1. (Top panel)**. Case notifications of Influenza, RSV and COVID-19 detected in Australia during the 2022 winter season. National Notifiable Disease Surveillance System (NNDSS) fortnightly data for influenza positive cases in NSW (orange triangles) and Australia (orange circles), COVID-19 (NSW, green triangles, Australia green circles) and RSV (Australia, purple circles) during 2022 are denoted by line graphs. **Figure 1 (bottom panel)**. Local sequenced influenza A cases included in this study (bars) were typed as H1N1 (pink) and H3N2 (blue), or not typed (grey). Local and national influenza positive cases were separated on a fortnightly basis (excluding 17 April week ending). Cases with influenza A and RSV co-infection were denoted by a star symbol (□) for RT-PCR detection and a diamond symbol (♦) for sequencing confirmation of RSV infection. RSV-positive cases were only illustrated when detected by a single platform; concurrent detections were not shown. Both symbols are displayed within the respective stacked bar for influenza A typing. RSV became a nationally notifiable disease as of 1 July 2021. However, the notification numbers shown in this diagram do not represent a national picture, as these conditions are not yet notifiable in all states and territories.

The spatiotemporal patterns of influenza A cases among NSW local health districts during the study period (March to July 2022) were visualised using monthly maps of NSW (Supplementary Figure 2). The first cases of influenza A/H3N2 (March) and A/H1N1 (April) from the study population were both diagnosed in Western Sydney. At least one case of each subtype was subsequently reported for each of the five local health districts represented in the study population.

### Influenza A viral diversity in New South Wales and evidence of within-subtype reassortment

Within our dataset, influenza A/H3N2 predominated across all timepoints after 10 April 2022 (79%, 103 of 130 genomes), concordant with state and national reports (Figure 1). A minority (22%, 28 of 130 genomes) of the A/H1N1 population (WHO clade 6B.1A.5a.2) also persisted throughout the season, with limited genetic diversity (Figure 2b). There was no difference in ages of cases infected with either subtype (p=0.232, paired two tailed t-test), and neither subtype was associated with hospitalisation at sampling (p=0.3416, paired two tailed t-test).

**Figure 2.**
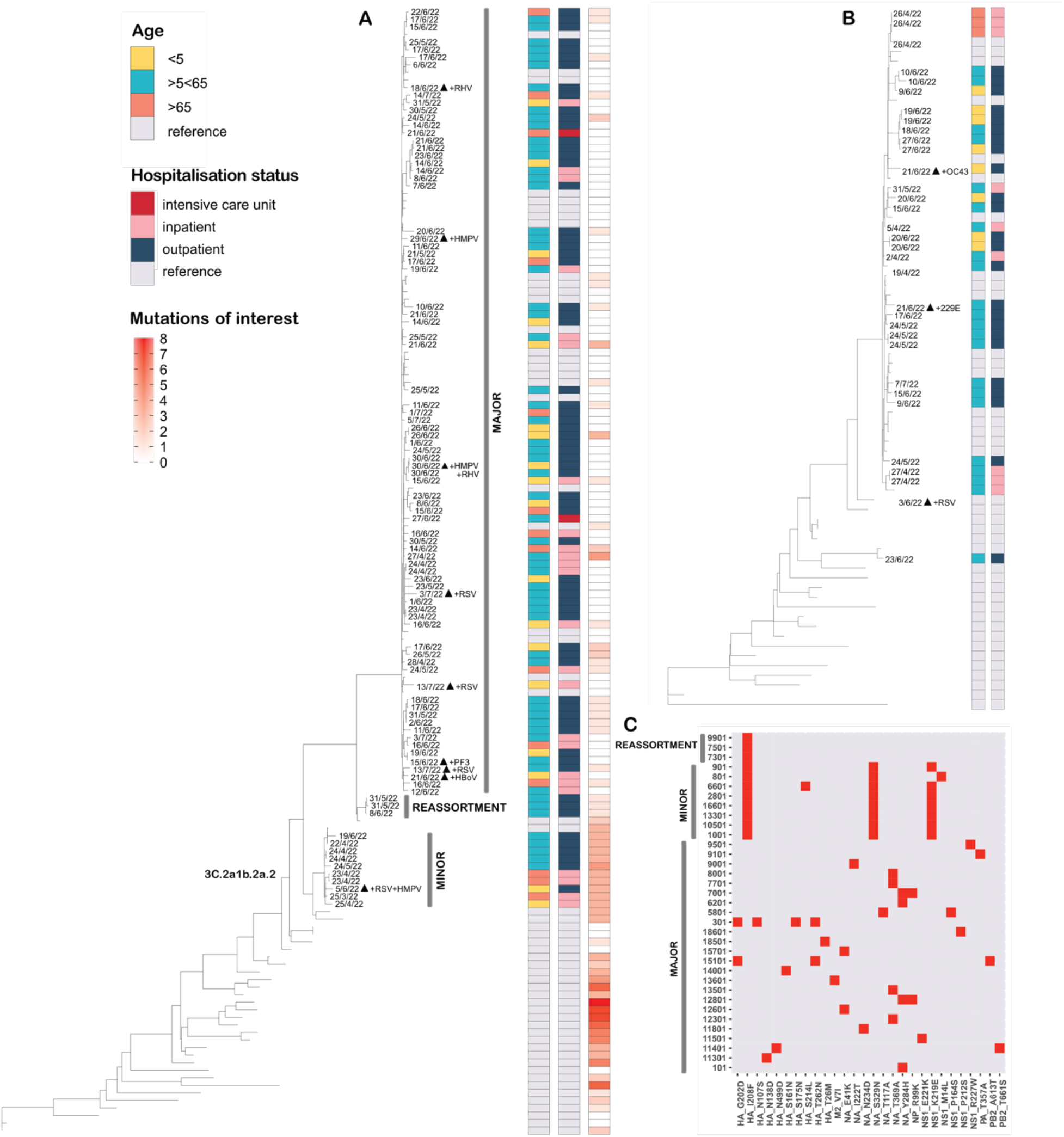
Influenza A virus phylogeny of circulating H3N2 and H1N1 clades during the Australian winter 2022. Influenza A virus phylogeny of circulating H3N2 (A) and H1N1 (B) cladesduring the Australian winter 2022. Age range of cases, hospitalisation status, and the number of mutations of high interest detected (A only) of each node are indicated on the colour bars. The date of collection and co-infection of each case sequenced in the study is labelled parallel to each tip. The major, minor and reassortant clades of A/H3N2 3C.2a1b.2a.2 are labelled in grey. C: the mutations of high interest as defined by Flusurver detected for A/H3N2 cases, with a higher frequency of mutations noted in the minor and reassortant clades.

Phylogenetic reconstruction of the A/H3N2 genomes, contextualised with reference genomes from the national WHO CC data, revealed the co-circulation of three distinct sub-clades of 3C.2a1b.2a.2, hereafter referred to as the “major” (87/100 H3N2 genomes, 87%), “minor” (10/100 genomes, 10%) and “reassortant” clades (3/100 genomes from this study, and one genome from the WHO CC) (Figure 2a).

Genomes belonging to the reassortant clade fell within the diversity of the major clade in segments PB2, PB1, NP, NA and NS, with complete nucleotide identity to several other genomes within this sub-clade. However, in segments PA, HA and MP, reassortant genomes clustered instead with the minor sub-clade (Supplementary Figure 4). The four reassortant genomes were obtained from samples collected in two distinct geographic regions (Metropolitan Greater Sydney, n=1, and rural Western NSW, n=3), over a period from 23 April – 8 June 2022, indicating sustained transmission of a single genetic lineage. These results suggest that the reassortant sub-clade arose from a single reassortment event, which brought together distinct circulating clades of the two major immunodominant regions of HA and NA with different recent ancestries.

We used FluSurver (35) to screen all genomes for presence of known phenotypically significant mutations that differed from the contemporaneous reference strain of the relevant subtype (H3N2 A/Darwin/6, and H1N1 A/Wisconsin/588). Mutations reported by FluSurver have been previously characterised to cause antigenic drift, change host specificity, antiviral susceptibility and/or affect glycosylation (Figure 2c). Genomes within the minor A/H3N2 clade contained four mutations of high interest within the HA (I208F and N112S), NA (S329N) and NS1 genes (K219E). All genomes from the minor and reassortant A/H3N2 clades contained both the HA mutations, I208F and N112S, in concordance with the evolutionary history of the reassortant HA segment (Supplementary Figure 4). Mutations HA I208F (HA1 192, classical H3N2 numbering) and HA N112S (HA1 96) are predicted to change host specificity and antibody recognition. Mutation I208F has been associated with adaptation in avian H5 influenza viruses by changing HA specificity from α2,6-to α2,3-linked sialic acid receptors (36). Mutation S329N (HA1 329) in the NA gene was detected in all genomes in the minor H3N2 clade. This mutation is predicted to generate an additional N-glycosylation site at position 329 which likely impacts the antigenic properties of the strain (37). All but one genome in the minor clade also contained mutation K219E (NS1 230) in the NS1 gene. NS1 blocks host cell antiviral responses through several mechanisms, and this mutation may contribute to increased virulence (38). No evidence of NA or endonuclease inhibitor resistance was identified.

To understand the subsequent evolutionary fate of the reassortant and the major and minor circulating A/H3N2 sub-clades, and to better understand the history of the individual segments, we explored global A/H3N2 sequences from the following Northern Hemisphere autumn/winter season.

### Influenza A/H3N2 genomic diversity between the Northern and Southern Hemisphere winters

We augmented our dataset with a similarly-sized set of 150 publicly available A/H3N2 whole genome sequences from international A/H3N2 cases sequenced between July 15^th^ and December 9^th^, 2022 (n=77 European, n=38 Northern America, n=19 Asia, n=7 South American, n=5 Middle East, n=4 Africa). We found that the A/H3N2 clades that dominated the 2022/2023 Northern Hemisphere winter were phylogenetically distinct from strains that circulated during the preceding Southern Hemisphere winter, as represented in our collection (Figure 3). Only seven genomes (n= 5 from Europe and n=2 from Asia) clustered with the major 3C.2a1b.2a.2 sub-clade circulating in Australia, and no genomes clustered with either the minor or the reassortant sub-clades.

**Figure 3.**
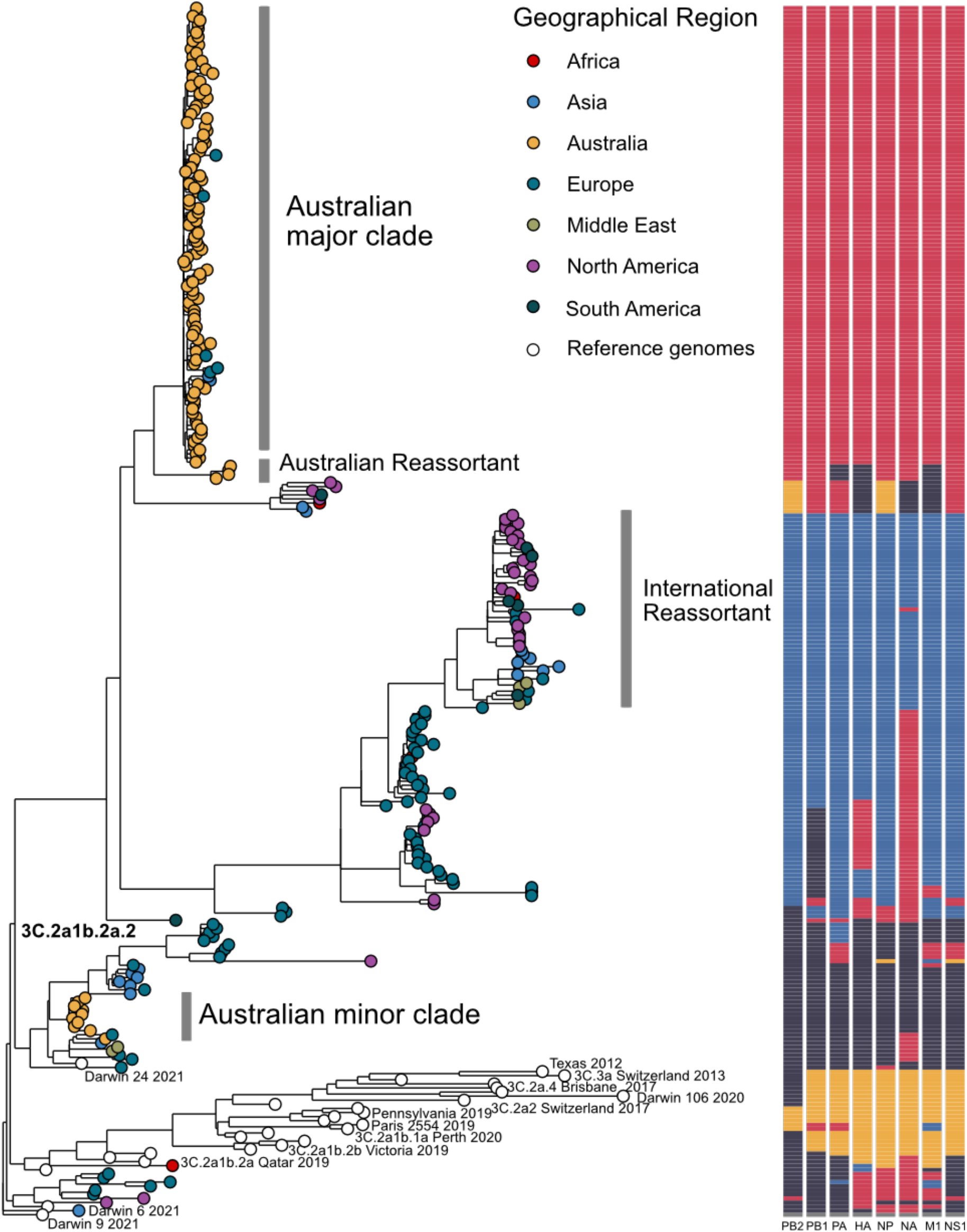
Influenza A/H3N2 whole genome phylogeny during the Northern and Southern hemisphere winters 2022. Influenza A/H3N2 WHO clade 3C.2a1b.2a.2 cases dominated the winter seasons in both the Northern and Southern Hemispheres during the winter 2022. Whole genome sequences recorded from A/H3N2 infections in Australia (orange nodes, n=103) were compared to international A/H3N2 genomes from Africa (red nodes, n=4), Asia (light blue nodes, n=19), Europe (dark green nodes, n=77), Middle East (grey nodes, n=5), northern (purple nodes, n=38) and southern (dark grey, n=7) America and historical reference genomes (white nodes, n=27). The major, minor and reassortant clades detected in Australia are annotated on the phylogeny. K-means clustering was performed on informative sites within each segment to visualise patterns of reassortment. The segment cluster for each genome is indicated by the colour bars. Segments are labelled below each bar indicating the Polymerase basic protein 2 (PB2), Polymerase basic protein 1 (PB1), RNA polymerase subunit (PA), hemagglutinin (HA), nucleoprotein (NP), neuraminidase (NA), matrix protein (M1) and non-structural protein (NS1).

Evidence of multiple instances of within-subtype reassortment could be readily inferred from phylogenetically incongruent clustering across the eight segments of the influenza A genome, just as was observed for the Australian reassortant clade (Figure 3). To visually represent this complex history for the global phylogeny of H3N2 clade 3C.2a1b.2a.2, we clustered all phylogenetically informative mutations and assigned an arbitrary colour to each cluster, for all eight segments. Every genome could thus be labelled with a collection of eight colour-coded histories for its eight segments, and the tree was “painted” in accordance with the evolutionary history of each genome (Figure 3). Clades which predominated in Europe shared sequence similarity at either NA and/or the HA segment with the major Australian sub-clade. Notable also were the nine clusters of putative reassortant clades, including the reassortant clade from the Australian season. These reassortants, with one exception, were confined to a single international geographic region. Interestingly the single reassortant clade which contains genomes from diverse geographical regions, contained a highly divergent NA segment which emerged in 1993 (Supplementary Figure 5). This demonstrates that selective sequencing of HA and NA genes limits the ability to differentiate globally-circulating strains and may mask the extent of diversification occurring through genome reassortment.

### Influenza A co-infection with respiratory viruses

In addition to the genomic epidemiology of influenza viruses post-COVID-19 restrictions, we were interested in the role of other common respiratory viruses circulating within the same population during the study period. One of the strengths of the capture-based sequencing approach is the ability to simultaneously sequence a diverse array of pathogens in the same assay, and we used this sequence data to look for the presence of co-infecting viruses in our study.

Among the 180 influenza A cases, 19 patients (10.6%) had respiratory viral co-infection identified by either qPCR (n=16), genome sequencing (n=20), or both (n=7) (Table 1, Supplementary Figure 3). Genomic sequencing uncovered 14 co-infecting viruses that had not been detected by routine RT-PCR respiratory virus assays. These include viruses which were not targeted by the RT-PCR assays used in our setting (human bocavirus HBoV [n=4], human coronavirus 229E [HCoV-229E; n=2] and HCoV OC43 [n=1]), not included in the testing requested for individual patients, or false negatives on routine RT-PCR screening (SARS-CoV-2 [n=3], HMPV [n= 4], RSV-A [n=1], RSV-B [n=4], human adenovirus hAdV [n=1]).

**Table 1.**
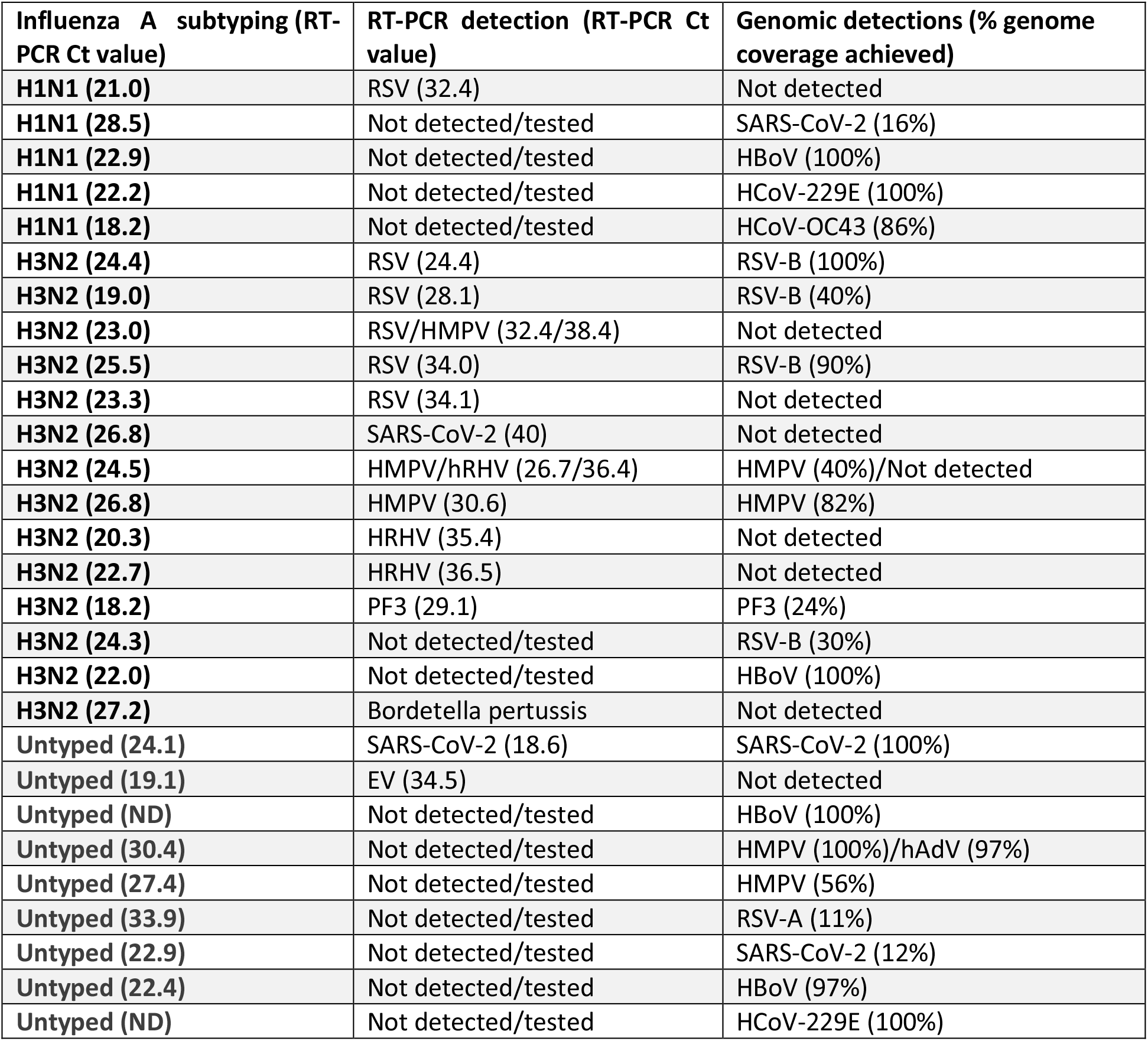
Comparison of respiratory virus co-infections detected by RT-PCR and genomics. Comparison of respiratory viral co-infection detection using genomics and RT-PCR. Different diagnostic respiratory testing algorithms were used by referring diagnostic laboratories during the study period, for example SARS-CoV-2 and Influenza testing were performed in some referring laboratories, whereas other included RSV, HMPV, RHV, PF, and EV. RSV, respiratory syncytial virus; HMPV, human metapneumovirus; RHV, rhinovirus; PF3, parainfluenza virus 3; SARS-CoV-2, severe acute respiratory syndrome 2; EV, enterovirus; HBoV, human bocavirus, HCoV-229E, human coronavirus 229E, HCoV-OC43, human coronavirus OC43, HRHV, human rhinovirus.

Complete or near complete genomes (>70% coverage) were obtained from 60% (12/20) of co-infecting pathogens, including HBoV (n=4/4), HCoV-229E (n=2/2), RSV-B (n=2/4), RSV-A (n=0/1), HMPV (n=2/4), HCoV-OC43 (n=1/1), SARS-CoV-2 (n=1/3) and hAdV (n=0/1).

Among our cohort, the most common respiratory viral co-infection with influenza A was RSV-B and HBoV (Table 1). Most patients with co-infections were within the 34–48-year-old age group (Supplementary Figure 3). Only one patient was confirmed positive for influenza A and two additional respiratory viruses (HMPV/HRV); this patient was within the 0–2-year-old age group (Supplementary Figure 3). In summary, we detected numerous and diverse co-infections with influenza A infections. Where multiple pathogens were detected and Ct values were available for both (n=14), most samples had large differences between the viral load of the co-detected pathogens, with median Ct difference (ΔCt) of 9.2 cycles (range 0 - 15.4, Table 1), consistent with consecutive rather than simultaneous infections, or with virus-virus interactions affecting the relative infection dynamics.

## Discussion

### Genomic epidemiology of influenza following easing of COVID-19 travel restrictions

During the Southern Hemisphere winter of 2022, numerous pathogens that cause symptomatic respiratory infections co-circulated in Australia after international travel resumed. International air travel for fully vaccinated residents resumed on 1 November 2021, travel between Australia states eased by March 2022 and fully vaccinated tourists were permitted to enter from 21 February 2022.(39-41). The rapid and early rise of influenza A cases in the autumn of 2022 raised concerns about low influenza vaccine uptake and effectiveness, importation and establishment of new influenza virus strains (by displacing current circulating influenza virus strains), increased disease severity due to co-infections, and the impact of co-circulating respiratory pathogens on healthcare resource utilisation. The season was dominated by circulation of A/H3N2 (3C.2a1b.2a.2) with persistent low prevalence of A/H1N1 infections. Indeed, the major A/H3N2 Australian clade reported in this study cause a spike in A/H3N2 in Victoria in autumn, this virus likely persisted and spread to other Australian states (24). A/H3N2 also dominated in the following Northern Hemisphere winter of 2022/2023, but from genetically distinct subclades of A/H3N2 3C.2a1b.2a.2.

Our findings demonstrated the added value of targeted enrichment such as RVOP, which allows the whole influenza viral genome to be sequenced in a single assay, in contrast to the traditional PCR-based methods for selective sequencing of only HA and NA genes. Our analyses of complete influenza genomes highlight evidence of gene segment reassortment within a single season, and a high prevalence of reassortment between Northern and Southern Hemisphere A/H3N2 genomes. A/H3N2 whole genome phylogenies showed a deep split between Australian and subsequently sampled global A/H3N2 viruses (Figure 3), suggesting that phylogenetically dissimilar clades caused A/H3N2 cases in the Northern Hemisphere., Despite their highly divergent backbones, a large proportion of the European and North American A/H3N2 genomes share similar NA and HA segments with the major clade circulating in Australia during its earlier winter season, as revealed by the phylogenetic structure of each segment (indicated by colour bars in Figure 3). In addition, up to nine reassortant A/H3N2 clades emerged, the vast majority of which were detected only within geographically distinct regions. This demonstrates the frequency and rapid diversification of influenza A viruses via within-subtype reassortment, which can be missed when only the HA and NA segments are sequenced.

We found no direct evidence of co-infection by distinct influenza viruses within a single sample, as expected given the limited sample size and previous reports indicating very low rates of dual influenza virus detection (42). However, the presence of multiple reassortments throughout the local and global A/H3N2 phylogeny strongly argues for relatively frequent and ongoing within-subtype co-infections, where multiple distinct lineages with each subtype co-circulate in the same population. Theses within-subtype co-infections would likely be masked by PCR based screen methods and maybe more apparent as genomic surveillance matures. The minimal phylogenetic divergence of the resulting reassortant lineages additionally supports the view that reassortment events do not require host shift or significant adaptation, and regularly produce evolutionarily fit viruses capable of sustained transmission within susceptible populations.(43, 44)

Examining influenza A, we assessed the extent to which the presence of other viruses, consistent with co-infection, was contributing to the patient load in our healthcare setting.(45, 46) We found that evidence of recent or concurrent infection by multiple pathogens, defined here as the relative proportion of sequence-detectable virus in symptomatic patients between 9 - 11% (16/180 and 19/180) had evidence of possible co-infection. This is comparable to previous studies that report co-infection rates of 14 - 15%(45, 47), where co-infections are more common in children less than 5 years. Of the sequence-confirmed possible influenza A co-infections, 5/19 (26%) were RSV (4/5 RSV B), consistent with the epidemic trend in Australia showing RSV to be co-circulating widely at the same time as the influenza A wave was peaking. Notably, no single virus dominated the co-infection profile with influenza A: we detected various viruses, including RSV, HMPV and HBoV, but also a variety of genotypes, with both RSV-A and RSV-B co-circulating. The predominant influenza A genotype was H3N2 (78% of samples), and correspondingly only 5 of 19 co-infections (26%) involved H1N1. The detection of RSV co-infection with A/H1N1 demonstrates that this is not a genotype-specific phenomenon, but rather reflects the epidemiological dynamics.

### Effectiveness of simultaneous multi-pathogen sequencing in public health

The method deployed in this study, the commercially available Illumina RVOP panel, was able to detect both RSV genotypes (A and B), with total RSV detection comparable with qPCR. However, this method may have been less sensitive to diverse enteroviral genomes such as rhinoviruses and it failed to recover human rhinovirus (HRV) from two samples where HRV was originally detected by PCR. This finding is consistent with similar reports of low sensitivity to enterovirus genomes by the competing Twist Comprehensive Viral Panel (48). In contrast, probe-based methods that use a denser set of probes to cover diverse viruses have substantially higher analytical sensitivity relative to the RVOP panel, and may be better suited to situations where monitoring for enteroviruses and other genetically diverse viral families is needed (18, 32).

The hybrid capture WGS-based approach is a powerful method and a useful intermediate tool between amplicon-based sequencing, which must be conducted through a separate laboratory workflow for each virus, and metagenomics, which is expensive and impractical for high throughput genomic epidemiology (49, 50). RVOP has the added advantage of multiple viral targets, remaining effective within regions of high genetic diversity, and permitting the detection of rapidly evolving, and potentially novel viruses (51-53). The utilisation of RVOP within our sequencing workflow provided the capacity to obtain high quality genomes of diverse influenza A strains circulating within the community, in addition to the detection of viral co-infections within our cohort.

Our study has limitations: this was a proof-of-concept deployment study in a public health setting, using a relatively limited number of remnant samples from routine microbiology testing, and thus does not explore the full capability of the methods. Research studies that used freshly collected samples and a custom probe panel designed to optimise sensitivity for genetically diverse pathogens (18, 32) demonstrate that improved performance is achievable in practice, with dedicated processes. Our sample numbers were also necessarily limited by available resources and timing, reducing power to detect all co-infections or those not circulating at the time. Our limits of detection were likewise conservative, and we considered only potential co-infections with sufficient viral load to return a positive sequencing result. However, we sampled representatively relative to the epidemic wave in our setting (Figure 1), and the close concordance between our results and national trends lend support to our conclusions. Unfortunately, influenza B which was at low levels of circulation during this study and was not detected in the cohort and therefore could not be assessed. In addition, antiviral resistance can be readily inferred from genomic data, importantly as we were able to generate complete influenza genomes, we had the capacity to infer resistance to NA and endonuclease inhibitors used to treat influenza infections, but neither were identified in our study.

Our study demonstrated that multi-pathogen genome sequencing can be beneficially deployed in a public health setting during epidemic waves of co-circulating respiratory viruses. This enables more efficient genomic epidemiological analysis and streamlines laboratory workflows, which currently are conducted independently for each individual virus species. We deployed a commercially available probe panel (RVOP, Illumina) to detect and sequence all commonly occurring respiratory viruses from clinical samples, using remnant material from routine microbiology testing. We demonstrated that in this setting, the sensitivity of probe panels is not inferior to routine RT-PCR in the diversity and multiplicity of respiratory viruses detected. Further refinement and customisation of such panels to increase probe density for genetically diverse pathogens could further enhance the sensitivity of this approach (32). Where practical, integration of testing workflows with sequencing would be expected to improve genome recovery and avoid the risk of RNA degradation inherent in the use of current sequential testing protocols. Expansion of sequences targeted by hybridisation panels have enabled identification, quantification of carriage load and typing of bacterial pathogens which may increase disease severity, such as *Streptococcus pneumoniae* and *Haemophilus influenzae* (54). Although increasing the number of probes adds to sequencing costs of capture-based sequencing methodologies, the ability to sequence a variety of pathogens, including coinfections, with a single assay could have significant cost advantages to public health laboratories. Costs can also be mitigated through workflow efficiencies and the development of new innovations to decrease costs per genome.

Further work is warranted to improve and expand the application of multi-pathogen genomics combined with syndromic surveillance, particularly in complex epidemic scenarios such as seasonal respiratory infections with enhanced surveillance of influenza subtypes to monitor for H5 and H7 infection in humans. Ongoing ripple effects from the COVID-19 pandemic are likely to continue to impact the dynamics of co-circulating pathogens and argue in favour of a shift towards multi-pathogen sequencing in communicable disease control.

## Data Availability

All data produced in the present study are available upon reasonable request to the authors. The Influenza A virus genome sequences and associated metadata can be found in the NCBI Sequence Read Archive online repository which can be accessed through (https://www.ncbi.nlm.nih.gov/sra/). Data pertaining to this study can be found in the BioProject PRJNA911331.

## Data availability

The *Influenza A virus* genome sequences and associated metadata can be found in the NCBI Sequence Read Archive online repository which can be accessed through (https://www.ncbi.nlm.nih.gov/sra/). Data pertaining to this study can be found in the BioProject PRJNA911331.

## Author contributions

Study concept and design by RR, TG. Sample processing and testing by CL, JA, WF, RR, SC. Sequencing, and analysis by RR, WF, TG, JA, SC, AA, CS. The first manuscript was written by JA, WF, RR, TG with additional editing from VS, JK, SC, DD, JA, SC, WF, SA, IB. The final manuscript was approved by all authors.

## Acknowledgements

This study was supported by the Virology Laboratory and Microbial Genomics Reference Laboratory at the Institute of Clinical Pathology and Medical Research-NSW Health Pathology. Referral of clinical samples from the NSW Health Pathology network of laboratories is appreciated. The authors would like to thank Sydney Informatics Hub, a Core Research Facility of the University of Sydney for providing research computing services.

## Methods

### Ethics

Governance and human ethics approval for clinical metadata and use of respiratory specimens from cases positive for *influenza A virus* in New South Wales (NSW), Australia were obtained by Western Sydney Local Health District Human Research Ethics Committee (2022/ETH02426).

### Samples for genomic surveillance

All clinical respiratory samples collected between March 25^th^ and July 14^th^, 2022 positive for *influenza A virus* by quantitative real-time polymerase chain reaction (RT-PCR) at the Institute of Clinical Pathology and Medical Research (ICPMR), Westmead and reference genomes provided by The WHO Collaborating Centre for Reference and Research on Influenza (WHO CCRRI) were included in the study.

### RNA extraction and RT-PCR

Total viral RNA was extracted using MagNA Pure 96 DNA and Viral NA Small Volume Kit (Roche Diagnostics Ltd., Rotkreuz, Switzerland), in accordance with the manufacturer’s instructions. An in-house, diagnostic multiplex TaqMan® probe real-time PCR panel (Supplementary Table 3) detecting the *influenza A virus* matrix gene was used. Extracts which had a RT-PCR detection of *influenza A virus* were subject to a second confirmatory RT-PCR to confirm RNA integrity prior to sequencing. The reaction was carried out in 20 μL volumes with 4 μL of template DNA with AgPath-ID™ One-Step RT-PCR Kit (Applied Biosystems™, ThermoFisher Scientific, Massachusetts, USA). The RT-PCR assays were conducted in a LightCycler® 480 System (Roche Diagnostics Ltd., Rotkreuz, Switzerland) using the following conditions: 45°C for 15 mins, 95°C for 15 mins, followed by 45 cycles of 95°C for 15 s and 60°C for 45 s, and 40°C for 30 s. Each run included a no template negative control (NTC).

### Targeted respiratory virus genome sequencing

Genomic sequencing was attempted on all 180 clinical extracts (Supplementary Figure 1). Viral enrichment was performed using the Illumina RNA Prep and Enrichment with the Respiratory Viral Oligo Panel version 2 (RVOP) (Illumina Inc., San Diego, California, USA). The RVOP comprehensive panel captures the genomes of 30 pathogenic human respiratory viruses, including SARS-CoV-2, and 12 common and recent *influenza A virus* subtypes (Supplementary Table 1). Total RNA extracts were used as input into the RNA Prep with Enrichment kit (Illumina Inc., California, USA). RNA denaturation, first and second strand cDNA synthesis, cDNA tagmentation, and library construction using unique dual indexes were performed according to manufacturer’s instructions. Individual libraries were purified then combined in equal volume 6-plex reactions for probe hybridisation. Probe hybridisation was performed overnight and held at 58°C, hybridised libraries were then captured and washed according to the manufacturer’s instructions and amplified as follows: initial denaturation 98 °C for 30 s, followed by 14 cycles of 98°C for 10 s, 60°C for 30 s, 72°C for 30 s, and a final extension of 72°C for 5 mins. Library pool quantities and fragment size were determined using Qubit™ 1X dsDNA HS Assay (Invitrogen™, ThermoFisher Scientific, Massachusetts, USA) and Agilent HS D5000 ScreenTape (Agilent Technologies Inc., California, USA), respectively. Resulting library captures were combined into an equimolar pool and sequenced using paired end 148 base pair reads on the Illumina MiniSeq or iSeq (Illumina Inc., California, USA) with the aim of generating 1×10^6^ raw reads per specimen (Figure 1).

### Bioinformatic processing

Demultiplexed sequence read pairs were classified by Kraken2 (55) using a custom database containing the human genome (GRCh38 build) and the full RefSeq set of bacterial and viral genomes. Sequences identified as either human or bacterial were removed using filter_keep_reads.py from the Castanet workflow (32). Remaining reads, composed of viral and unclassified reads, were trimmed to remove Illumina adapter sequences using Trimmomatic version 0.36 (56)], with the ILLUMINACLIP options set to “2:10:7:1:true MINLEN:50” to retain only high quality reads. Trimmed reads were mapped to the influenza A virus genome of isolates A/Wisconsin/588/2019 [EPI_ISL_404460] for H1N1 and A/Darwin/9/2021 [EPI_ISL_16998754] for H3N2, using shiver version 1.5.7 (57), with bowtie2 (58) as the mapper. Only properly paired reads with insert size <200-bp and with at least 70% sequence identity to the reference were retained. Consensus calls were derived for positions supported by a minimum of 10 mapped reads.

### Phylogenetic analysis

Genomes were included in phylogenetic analyses if the consensus sequence consisted of no more than 25% ambiguous characters. To place these data into the global phylogenetic context and to help resolve ancestry, a collection of consensus sequences for A/H1N1 and A/H3N2 viruses were downloaded from the GISAID database (59) on December 15^th^, 2022, and included in the set of sequences to be aligned. Sequences included ancestral reference genomes as well as all genomes spanning the period between the end of the Southern Hemisphere winter and the start of the Northern Hemisphere winter season (August-December 2022). All available complete genomes for H1N1 and H3N2 were downloaded, and manually curated to retain only sequences listed as “original specimen” to remove *in vitro-*generated viral diversity. Reference genomes from the 2022 global collection were downsampled using epidemiologically weighted random sampling, with each genome given a weight 1/x_*i*_, where x_*i*_ is the number of sequences from location *i*, for each of A/H3N2 and A/H1N1. This procedure reduced the impact of oversampled locations while retaining maximal geographic representation. The resulting reference genomes, together with sequences from this study, were aligned using MAFFT (60) to produce one alignment each for H1N1 and H3N2. For gene-specific analyses, the regions corresponding to each segment were selected from the whole-genome alignment.

Maximum-likelihood phylogenetic reconstruction was performed separately on the whole-genome alignments and all eight gene segment alignments, with automated model selection as implemented in IQ-TREE (61), and time resolved phylogenies were produced using TreeTime (62). A total of 1000 bootstrap replicates were used for each IQ-TREE run, and zero-length branches were collapsed. For tree reconstruction, the first 7 codons (21 bases) of each segment were masked to prevent spurious phylogenetic signal arising from ambiguous reporting of segment ends due to different primer-based amplification protocols used in the production of global reference sequences.

### RSV genotype classification

RSV genotypes A and B were identified by sequence similarity to representative consensus genomic sequences of RSV-A and RSV-B, generated as follows: complete RSV-A (n=1295) and RSV-B (n=834) genomes were downloaded from NCBI Virus (https://www.ncbi.nlm.nih.gov/labs/virus/vssi) and reduced to a set of representative sequences at 90% sequence identity using cd-hit-est (63) with cluster threshold 0.9, generating four clusters for RSV-A and 3 for RSV-B. Similarity to RSV-A and RSV-B consensus sequences was used for classification.

### Phylogenetic clustering of whole genomes of influenza A/H3N2

To visually represent the structure of the A/H3N2 whole genome phylogeny, we implemented a simplified procedure for phylogeny “painting” conceptually similar to ChromoPainter and fineSTRUCTURE (64): specifically, for each genomic segment, we painted the tree in four arbitrary colours corresponding to the most highly informative splits in the phylogeny, using informative sites within that segment to identify the splits. Phylogenetically informative sites (SNPs) were clustered using the k-means algorithm as implemented in the Python library SciPy (v12.0), testing values of k from 2 to 8 for the number of clusters. The optimal concordance with the deep structure of the whole-genome phylogeny was obtained with k=4, that is, the tree structure could be “painted” using four colours. The predicted clusters for each segment were aligned to the corresponding node on the whole genome phylogeny using ggtree (65).

