## Supplementary Material for "Respiratory virus genomic epidemiology during post-pandemic re-emergence of influenza in Australia"

**Supplementary Figure 1**. Outline of RT-PCR positive NAAT Influenza specimens used in this study.

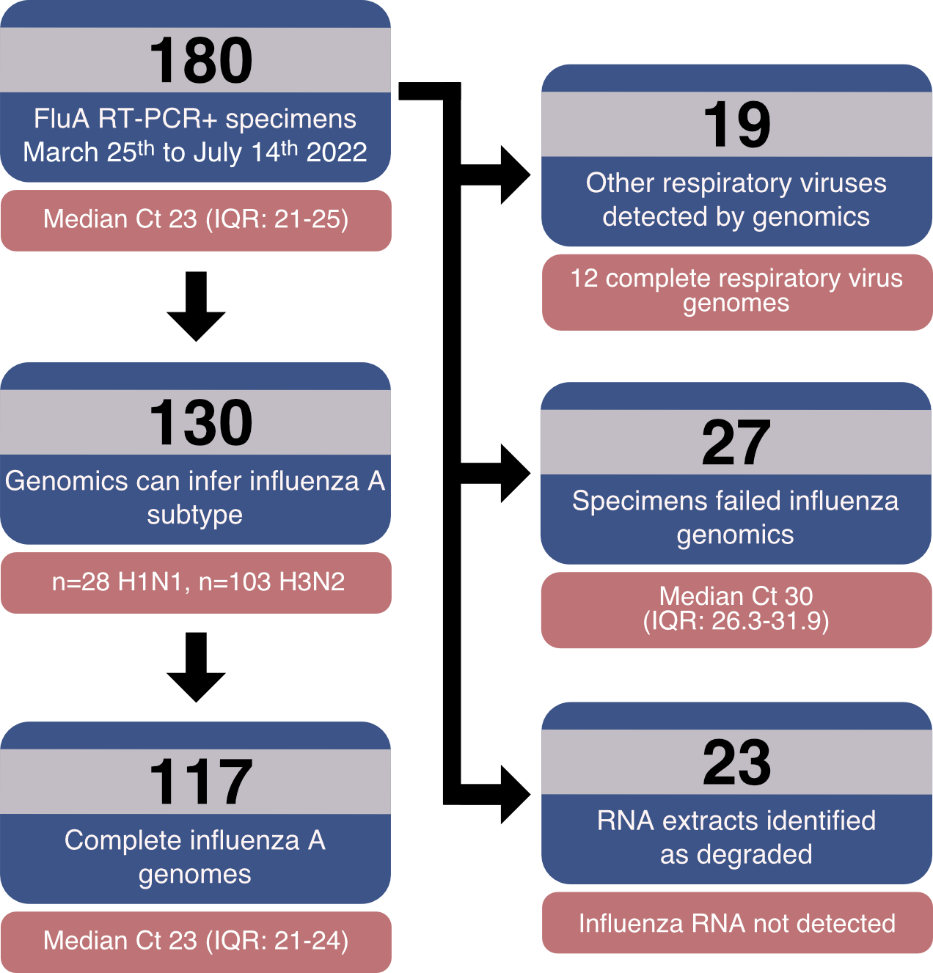

*Cycle Threshold value (Ct), Interquartile range (IQR)

Supplementary Figure 1. Successful genome reconstruction was highly concordant with the amount of influenza RNA template, consistent with previous reports that targeted capture protocols such as RVOP are quantitative with respect to viral load. Specimens from which a complete genome was obtained had an average cycle threshold value of 22.5 (interquartile range 20.7 – 23.8, n=117) whereas specimens from which we could not reconstruct whole genomes had an average cycle threshold value of 30.2 (interquartile range 26.3 – 31.9), including 23 specimens where Influenza could not be detected by RT-PCR prior to sequencing, consistent with RNA degradation in these samples.

**Supplementary Figure 2**. Geographic patterns of Influenza infection in NSW during the study period.

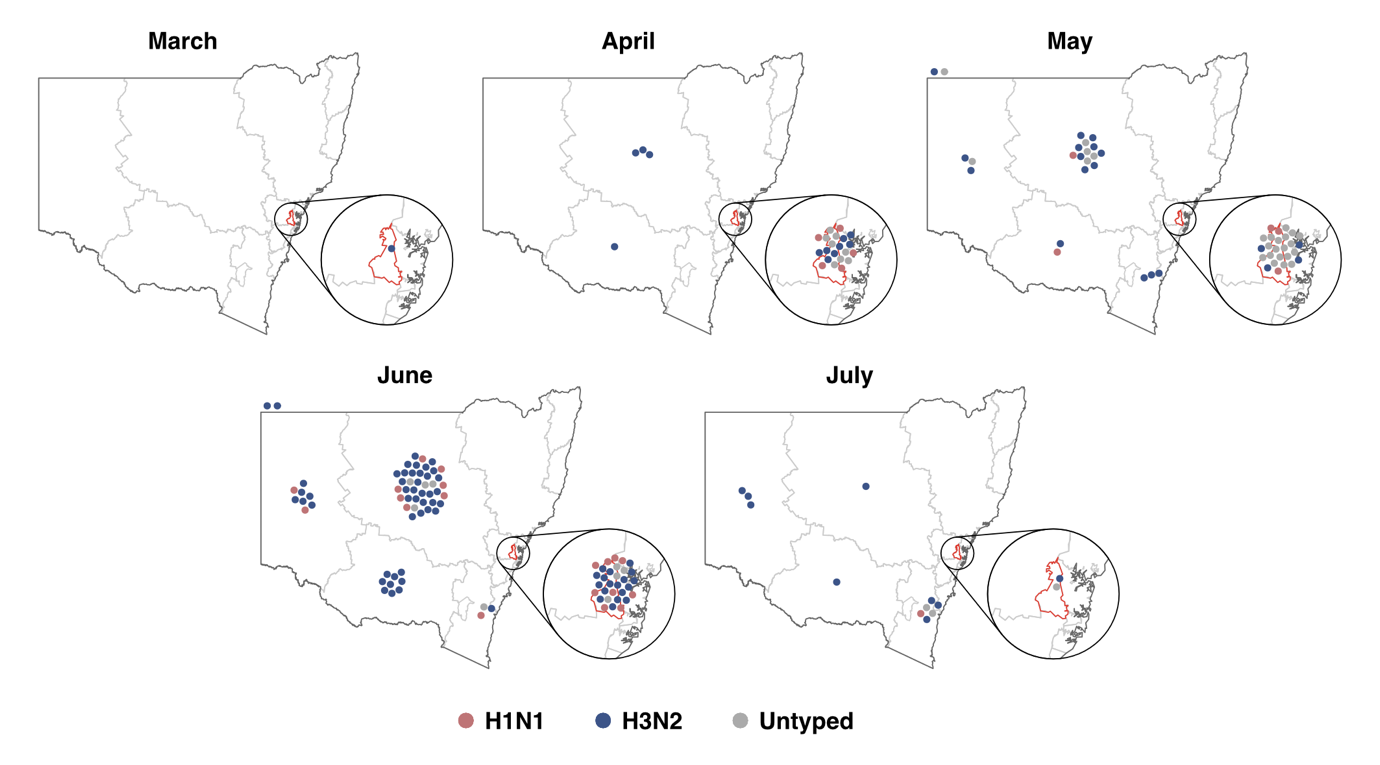

**Supplementary Figure 2.** Geographical distribution of confirmed influenza A cases in New South Wales from March to July 2022. Viruses were subtyped as H1N1 (pink) and H3N2 (blue) or were not typed (grey). Each filled circle represents a single PCR confirmed influenza A case. New South Wales was divided into NSW Health local health districts (grey lines) covering the Sydney metropolitan region (n=8), and rural and regional areas (n=7). The expanded portion (black circle) in each month represents the number of PCR confirmed cases within the Western Sydney Local Health District. Cases located above the map (n=4) were not assigned a local health district due to missing metadata. The placement of circles is not representative of the location of patient sample collection: they are placed near the centre of the corresponding local health district (LHD).

**Supplementary Figure 3.** Age range of PCR confirmed influenza A cases by admission status and respiratory co-infection of confirmed influenza A cases in New South Wales from March to July 2022

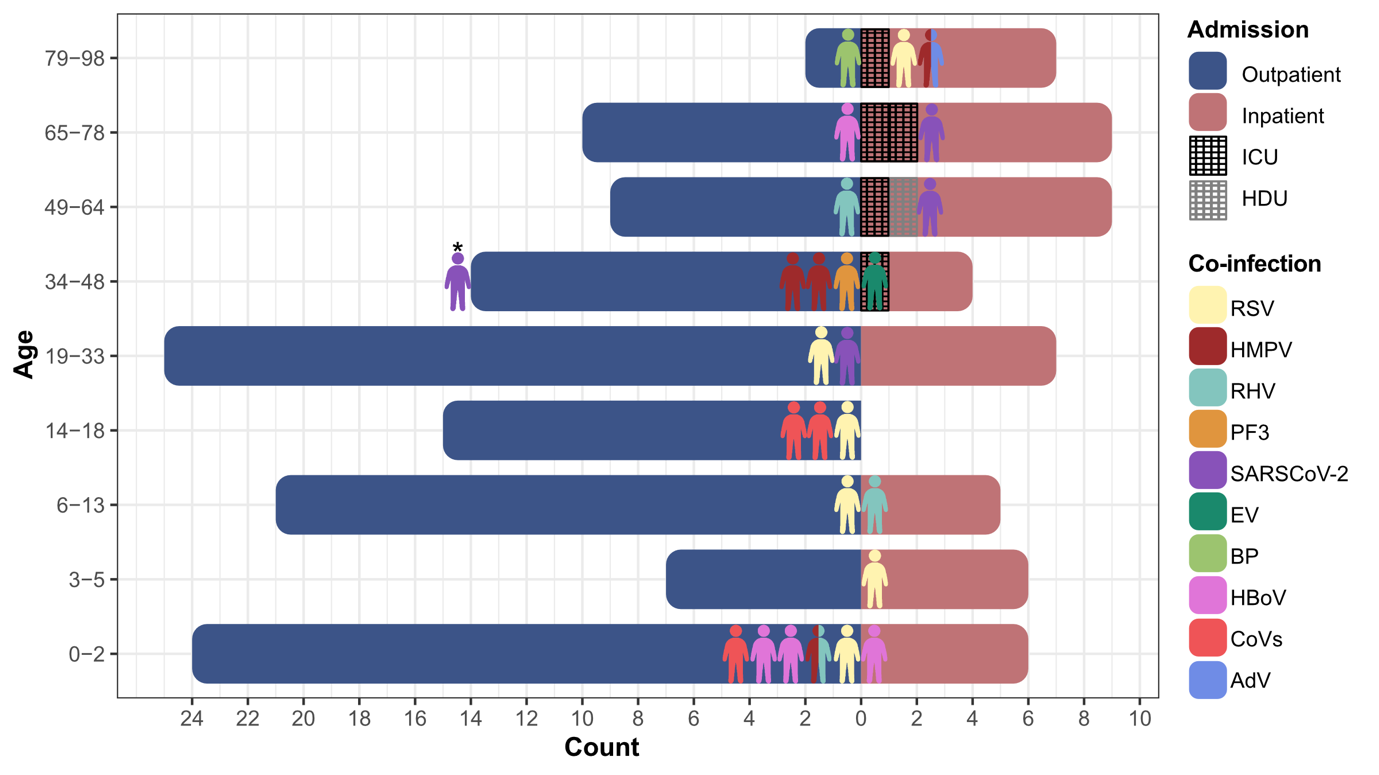

**Supplementary Figure 3:** Population pyramid for age of PCR confirmed influenza A cases by admission status and respiratory co-infection. Admission of positive cases was classified as either outpatient and not admitted to hospital (blue), or inpatient and admitted to hospital (pink). Inpatients were further classified as admission to units of critical care, including ICU (black hatch) or HDU (grey hatch). Patients with respiratory co-infections confirmed by RT-PCR and/or sequencing were denoted by coloured silhouettes, yellow (RSV, respiratory syncytial virus, n=6), red (HMPV, human metapneumovirus, n=4), light blue (rhinovirus, RHV, n=3), orange (PF3, parainfluenza virus 3, n=1), purple (SARS-2, severe acute respiratory syndrome coronavirus 2, n=3), dark green (EV, enterovirus, n=1), light green (BP, *Bordetella pertussis,* n=1), pink (HBoV, human bocavirus, n=4), red (CoVs, human coronaviruses OC43 (n=1), 229E (n=2)) and human adenovirus (n=1). *Hospitalisation status unknown.

**Supplementary Figure 4.** Time resolved phylogenies of Influenza A/H3N2 genome segments

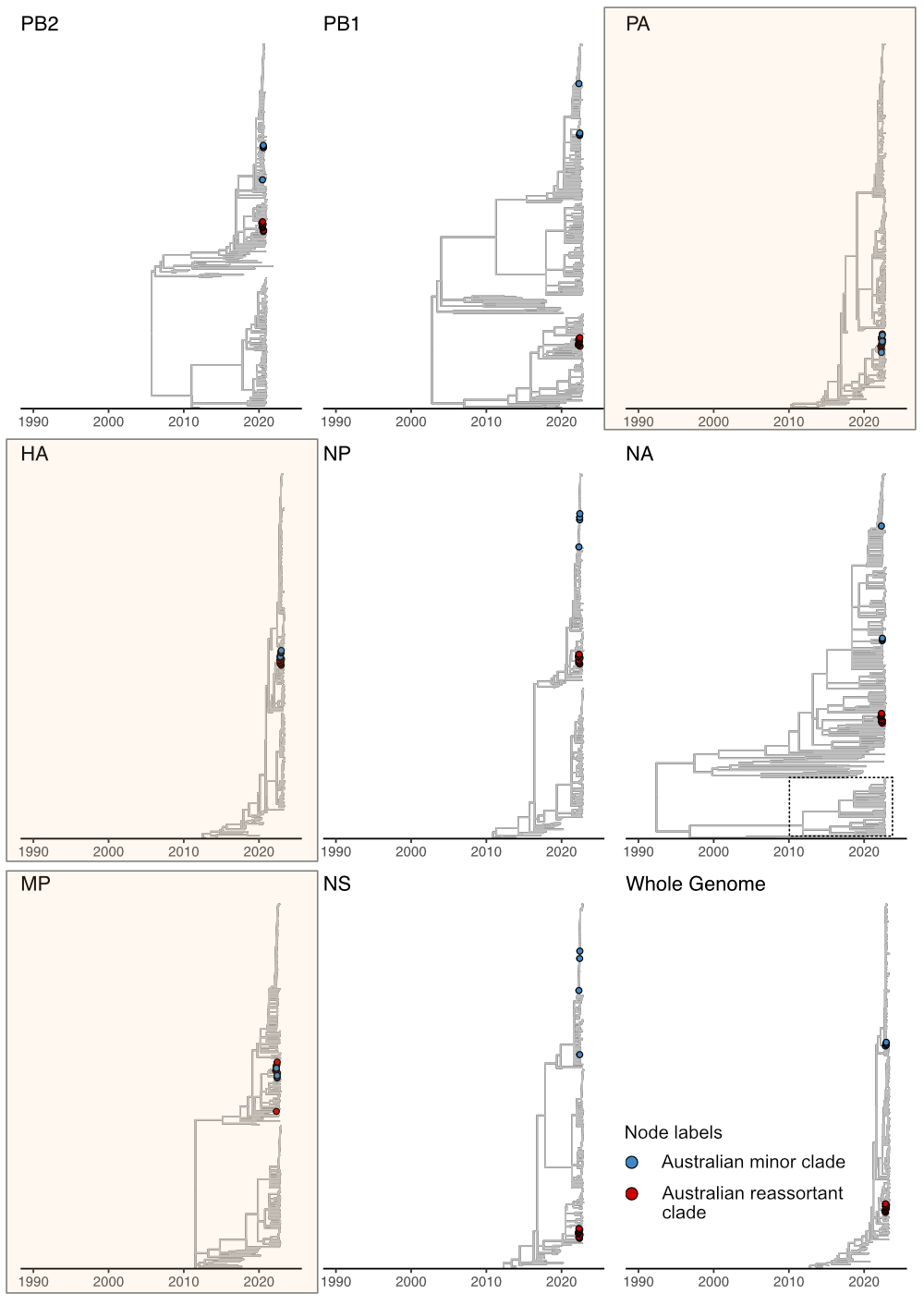

**Supplementary Figure 4.** Influenza A/H3N2 time resolved segment phylogenies. Whole genome sequences and individual segment phylogenies were time resolved to investigate the evolutionary history of the Australian minor and recombinant clades. Blue nodes indicate genomes from the Australian minor clade and red nodes indicate genomes from the Australian reassortant clade. In contrast to all other segments the Australian minor and reassortant clade share evolutionary history in the PA, HA and MP segments (orange shaded background). The highly divergent neuraminidase (NA) segment shared by all genomes in the international reassortant clade is indicated by the dashed box. Polymerase basic protein 2 (PB2), Polymerase basic protein 1 (PB1), RNA polymerase subunit (PA), hemagglutinin (HA), nucleoprotein (NP), neuraminidase (NA), matrix protein (M1) and non-structural protein (NS1).

**Supplementary Table 1: Bioinformatic quality metrics of Influenza genomes and co-infections generated in this study**

**Supplementary Table 2:** Viruses targeted by Illumina’s Respiratory Viral Oligo Panel (RVOP) target enrichment sequencing workflow. Viruses were classified by viral family; single-stranded (*Orthomyxoviridae*, n=13; *Picornaviridae*, n=7; *Coronaviridae*, n=5; *Paramyxoviridae*, n=4; *Parvoviridae*, n=4; *Pneumoviridae*, n=3), and double-stranded (*Adenoviridae*, n=3; *Polyomaviridae*, n=2).

| **Virus** | **Classification** | **Virus** | **Classification** |
| --- | --- | --- | --- |
| Human coronavirus 229E | + ss *Coronaviridae* | *Influenza A virus* (A/ Texas/50/2012(H3N2)) | – ss *Orthomyxoviridae* |
| Human coronavirus NL63 | + ss *Coronaviridae* | *Influenza A virus* (A/ Michigan/45/2015(H1N1)) | – ss *Orthomyxoviridae* |
| Human coronavirus OC43 | + ss *Coronaviridae* | *Influenza B virus* (B/Lee/1940) | – ss  *Orthomyxoviridae* |
| Human coronavirus HKU1 | + ss *Coronaviridae* | *Influenza B virus* (B/ Wisconsin/01/2010) | – ss *Orthomyxoviridae* |
| SARS-CoV-2 | + ss *Coronaviridae* | *Influenza B virus* (B/ Brisbane/60/2008) | – ss *Orthomyxoviridae* |
| Human parainfluenza virus 1 | – ss *Paramyxoviridae* | *Influenza B virus* (B/ Colorado/06/2017) | *Orthomyxoviridae* |
| Human parainfluenza virus 2 | – ss *Paramyxoviridae* | *Influenza B virus* (B/ Washington/02/2019) | – ss *Orthomyxoviridae* |
| Human parainfluenza virus 3 | – ss *Paramyxoviridae* | *Human bocavirus 1* (Primate bocaparvovirus 1 isolate st2) | – and + ss *Parvoviridae* |
| Human parainfluenza virus 4a | – ss *Paramyxoviridae* | *Human bocavirus 2c* PK isolate PK-5510 | – and + ss *Parvoviridae* |
| Human metapneumovirus (CAN97-83) | – ss *Pneumoviridae* | *Human bocavirus 3* | – and + ss *Parvoviridae* |
| Respiratory syncytial virus (type A) | – ss *Pneumoviridae* | Human bocavirus 4 NI strain HBoV4- NI-385 | – and + ss *Parvoviridae* |
| Human Respiratory syncytial virus 9320 (type B) | – ss *Pneumoviridae* | KI polyomavirus Stockholm 60 | ds *Polyomaviridae* |
| Human adenovirus B1 | ds *Adenoviridae* | WU Polyomavirus | ds *Polyomaviridae* |
| Human adenovirus C2 | ds *Adenoviridae* | Human parechovirus type 1 PicoBank/HPeV1/a | + ss *Picornaviridae* |
| Human adenovirus E4 | ds *Adenoviridae* | Human parechovirus 6 | + ss *Picornaviridae* |
| Influenza A virus (A/Puerto Rico/8/1934(H1N1)) | – ss *Orthomyxoviridae* | Human rhinovirus A89 | + ss i |
| Influenza A virus (A/ Korea/426/1968(H2N2)) | – ss *Orthomyxoviridae* | Human rhinovirus C (strain 024) | + ss *Picornaviridae* |
| Influenza A virus (A/New York/392/2004(H3N2)) | – ss *Orthomyxoviridae* | Human rhinovirus B14 | + ss *Picornaviridae* |
| Influenza A virus (A/goose/ Guangdong/1/1996(H5N1)) | – ss *Orthomyxoviridae* | Human enterovirus C104 strain: AK11 | + ss *Picornaviridae* |
| Influenza A virus (A/Zhejiang/DTIDZJU01/2013(H7N9)) | – ss *Orthomyxoviridae* | Human enterovirus C109 isolate NICA08-4327 | + ss *Picornaviridae* |
| *Influenza A virus* (A/Hong Kong/1073/99(H9N2)) | – ss *Orthomyxoviridae* | Human control genes |  |

**Supplementary Table 3:** Real-time PCR primer and panels utilised in-house for the detection of viral respiratory pathogens.

| **Target Virus** | **Gene** | **Strand** | **Primers** |
| --- | --- | --- | --- |
| *Influenza A virus* | Matrix | Forward | 5'–GAC CAA TYY TGT CAC CTY TGA C–3' |
|  |  | Reverse | 5'–FGG GCA TTY TGR ACA AAD CGT CTA–3' |
|  |  | Probe | 5'–TxRd–CGT GCC CAG TGA GCG RGG ACT GCA–BHQ2–3' |
| *Influenza B virus* | NP | Forward | 5'–CAACGATGACATGGAGAGRAAC–3' |
|  |  | Reverse | 5'–GCCTCCTGTTTTGTTGTGATC–3' |
|  |  | Probe | 5'–Q670–CCTTCTTTSACATCTCTGGCATTCTT–BHQ2–3' |
| *Human orthopneumovirus* (RSV) | NP | Forward | 5'–TAG TGT RCA RGC AGA AAT GG–3' |
|  |  | Reverse | 5'–AGT GRG GAA ATT GAG TCA AAG AT–3'  5'–RGG RAA TTG AGT TAA TGA CAG C–3' |
|  |  | Probe | 5'–FAM–CGT GCC CAG TGA GCG RGG ACT GCA–BHQ1–3' |
| Rhinovirus | ‘5 UTR | Forward | 5'–GCC CCT GAA TGY GGC TAA–3' |
|  |  | Reverse | 5'–GAA ACA CGG ACA CCC AAA GTA–3' |
|  |  | Probe | 5'–FAM–TGG TCC CRT CCC GCA MTT GC–BHQ1–3' |
| *Human respirovirus 3* (parainfluenza) | Matrix | Forward | 5'–GAA GTG AGA AGA ACA GTY AAA GC–3' |
|  |  | Reverse | 5'–CAT TGA GGA GCA AGA GCA AC–3' |
|  |  | Probe | 5'–FAM–TTG GCA TCG AAC ARC ATT CC–BHQ1–3' |
| *Enterovirus* |  | Forward | 5'–GCC CCT GAA TGY GGC TAA–3' |
|  |  | Reverse | 5'–GAA ACA CGG ACA CCC AAA GTA–3' |
|  |  | Probe | 5'–Red610–CGG TTC CGC TGC RGA GTT RGC C–BHQ2–3' |
| Human adenovirus | Hexon | Forward | 5'–BCA GGA YGC CTC GGA RTA–3' |
|  |  | Reverse | 5'–AAA CTT GTT XYY CAG GST GAA GTA SGT–3' |
|  |  | Probe | 5'–FAM–ART TYG CCC GYG CCA CSG–BHQ1–3' |
| *Human metapneumovirus* | NP | Forward | 5'–CAT AYA ARC ATG CTA TAT TAA AAG AGT CTC–3' |
|  |  | Reverse | 5'–CCT ATY TCT GCA GCA TAT TTG TAA TCA G–3' |
|  |  | Probe | 5'–Q670–TCT TGY TGC AAT GAT GAR GGT GTY AC–BHQ2–3' |

NP, nucleoprotein gene; TxRd, Texas Red fluorophore; Q670, Quasar 670 fluorophore; FAM, fluorescein amidites fluorophore; Red610.
